# The Impact of School Closure for COVID-19 on the US Healthcare Workforce and the Net Mortality Effects

**DOI:** 10.1101/2020.03.09.20033415

**Authors:** Jude Bayham, Eli P. Fenichel

## Abstract

**Background:** COVID-19 is leading to the implementation of social distancing policies around the world and in the United States, including school closures. The evidence that mandatory school closures reduce cases and ultimately mortality mostly comes from experience with influenza or from models that do not include the impact of school closure on the healthcare labor supply or the role of the healthcare labor force in reducing the per infection mortality from the pathogen. There is considerable uncertainty of the incremental effect of school closures on transmission and lives saved from school closures. The likely, but uncertain, benefits from school closure need to be weighed against uncertain, and seldom quantified, costs of healthcare worker absenteeism associated with additional child care obligations.

**Methods:** We analyze data from the US Current Population Survey to measure the potential child care obligations for US healthcare workers that will need to be addressed if school closures are employed as a social distancing measure. We account for the occupation within the healthcare sector, state, and household structure to identify the segments of the healthcare labor force that are most exposed to child care obligations from school closures. We use these estimates to identify the critical level for the importance of healthcare labor supply in increasing a patient’s COVID-19 survival probability that would undo the benefits of school closures and ultimately increase cumulative mortality.

**Findings:** The US healthcare sector has some of the highest child care obligations in the United States. 29% of healthcare provider households must provide care for children 3-12. Assuming non-working adults or a sibling 13 years old or older can provide child care, leaves 15% of healthcare provider households in need of childcare during a school closure, while 7% of healthcare households are single-parent households. We document the substantial variation within the healthcare system. For example, 35% of medical assistants and 31% of nursing, psychiatric, and home health aide households have child care obligations, while only 24% of emergency medical personnel have childcare obligations. Child care obligations can vary between states by over 10 percentage points. A 15% decline in the healthcare labor force, combined with reasonable parameters for COVID-19 such as a 15% case reduction from school closings and 2% baseline mortality rate implies that a 15% loss in the healthcare labor force must decrease the survival probability per percent healthcare worker lost by 17.6% for a school closure to increase cumulative mortality. This means that the per infection mortality rate cannot increase from 2% to 2.35% when the healthcare workforce declines by 15%; otherwise, school closures will lead to a greater number of deaths than they prevent. For school closures to unambiguously provide a net reduction in COVID-19 mortality with these parameters, the school closures must reduce cases by over 25%.

**Conclusion:** School closures come with many tradeoffs. Setting aside economic costs, school closures implemented to reduce COVID-19 spread create unintended childcare obligations, which are particularly large in healthcare occupations. Detailed data are provided to help public health officials make informed decisions about the tradeoffs associated with closing schools. The results suggest that it is unclear if the potential contagion prevention from school closures justifies the potential loss of healthcare workers from the standpoint of reducing cummulative mortality.

**Funding:** No external funding.

**Research in Context:** *Evidence before this study:* COVID-19 is affecting many countries around the world. Multiple countries, states, and school districts are employing school closures as a non-pharmaceutical social distancing strategy. Support for school closures mostly comes from models and experience with influenza. Few studies explicitly consider the tradeoff between case reduction and disease burden with the potential loss of healthcare workers to child care obligations. We found only two studies that attempted to quantify the potential child care burden of school closures for healthcare workers. These analyses were coarse, and no studies have explicitly considered the tradeoff between reduced transmission and healthcare labor’s role in cumulative mortality.

*Added value of this study:* Using current detailed data from the US Current Population Survey, we quantify the exposure of the US healthcare sector, occupations within the healthcare sector, and individual US states to unmet child care obligations for US healthcare workers in the event of a school closure. These obligations could compromise the ability of the US healthcare system to respond to COVID-19. We identify the parameter space where school closure could lead to a greater number of deaths from COVID-19 through healthcare labor force reductions. We find that the current best estimates of healthcare worker likely absenteeism to provide child care in the event of school closures imply great uncertainty to whether school closures will ultimately reduce COVID-19 mortality.

*Implication of all the available evidence:* Targeted pharmaceutical interventions for COVID-19 are likely months away. However, supportive measures by healthcare providers are important. Social distancing, including school closures, can limit COVID-19 cases. However, the evidence that the potential transmission reduction benefits of mandatory school closures exceed the costs of potentially imposing greater child care obligations on healthcare workers, thereby reducing the healthcare workforce, is not overwhelming. We provide the first explicit analysis of the school closure tradeoff between case reduction and labor force impact on patient survival probability. A 15% decline in the healthcare labor force, combined with reasonable parameters for COVID-19 such as a 15% case reduction from school closings and 2% baseline mortality rate implies that a 15% loss in the healthcare labor force must decrease the survival probability per percent healthcare worker loss by 17.6% for school closures to increase cumulative mortality. This means that the per infection probability of survival cannot increase by more than 2.4%; otherwise, school closures will lead reduce patient survival. Even if the only objective is to save lives, there is still a tradeoff associated with closing schools due to potential losses in healthcare labor force capacity.

## Introduction

Covid-19’s global spread is triggering a range of public health responses. School closures are some of the highest-profile social distancing measures employed to slow the spread of an infectious disease. Japan recently instituted a nationwide school closure, which drew a swift response from families scrambling to make alternative childcare arrangements. South Korea, Italy, and some US school districts have closed schools. School closures prevent contacts among children and reduce cases. But, there is a downside to closing schools, even if the only goal is saving lives during an epidemic. Closing schools can inadvertently cause child care shortages that strain the healthcare system. Lempel et al. estimated that the child care obligations associated with school closure could reduce key medical personnel by 6-19%.^1^ Here, we use the recent years (2018-2020) of the US Current Population Survey to estimate the school closure induced child care obligations for the US healthcare labor force. Approximately 29% of US households with a healthcare worker have at least one school-aged child age 3-12, who requires child care. Child care obligations are especially high and relatively inflexible in households where a parent is a nurse or medical assistant. Understanding these tradeoffs are important in planning the public health response to COVID-19, because if the survival of infected patients is sufficiently sensitive to declines in the healthcare labor force, then school closures could increase deaths.

The benefit from closing schools during an epidemic is to reduce transmission and new cases. Cauchemez et al. estimated that extended school closures in France could reduce H5N1 influenza cases by 13-17%.^2^ Bayham et al. show that schools are likely places for transmission among US children.^3^ However, Bayham et al. also show voluntary behavioral changes, without mandatory shutdowns, appeared to reduce cases of the 2009 H1N1 influenza on the order of 10-13%. In a systematic review, focused on influenza and school closures, Jackson et al. find some evidence that school closures are effective, but that the empirical evidence does not resolve how or when to close schools.^4^ Furthermore, they find school closure mostly reduces infection in school children. Koh et al. do not find strong evidence that school closures prevent the spread of hand, foot, and mouth disease.^5^ Adda finds that while school closures do reduce incidence in France, the economic costs are large.^6^ The benefits of school closure are often estimated relative to a baseline of no voluntary changes in behavior, but it is likely that the correct baseline for forcasting the school closure effect on COVID-19 includes other voluntary behavioral changes.

The potential benefits of school closures must be balanced with the costs. Smith et al., Lempel, Epstein, and Hammond, and Adda analyze the economic impacts of school closures.^1,6,7^ Schooling is one of the most important investments we make in our children’s futures, and we do not have good estimates of how prolonged school closures influence drop-out rates and future earnings. This uncertainty makes a holistic assessment of tradeoffs challenging.

Setting aside the economic costs and focusing on reducing mortality, school closures still create a tradeoff. Many healthcare workers must reduce time spent providing patient care, running diagnostic tests, and tracing contacts, and increase time caring for their own children. This tradeoff should not be ignored because the capacity to care for infected individuals and trace contacts directly influences the development of an epidemic, the survival of infected patients, and ultimately cummulative mortality from the pathogen.

## Methods

First, we highlight two pathways school closure can effect pathogen induced mortality. School closures, *c*, can affect mortality via (1) reduction in cases and (2) a reduction in the healthcare labor force that treats sick patients and prevents mortality. While no pharmaceutical treatment is available, supportive measures are still important for patient survival. Define cumulative mortality as

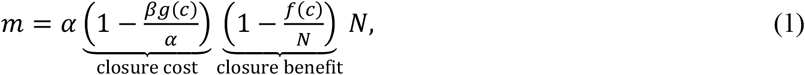

where *α* is the baseline mortality fraction for *N* cases, *-βg(c)/α* is the percent increase in the case mortality rate through the reduction in healthcare labor force from a school closure (*g(c) < 0*), and *f(c)/N* is the percent decrease in cumulative cases from the school closure. If there are no school closures, *c* = 0, then 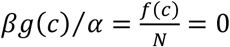. This implies mortality is *m= αN*. This model highlights the tradeoff between the case-reducing effect of school closures and the cost in terms of lost healthcare labor supply.

Analyzing the net effect of school closures on mortality requires estimation of three factors that are not part of canonical epidemiological models.^8,9^ The first term is *g(c)*, which is the effect of school closure on the healthcare labor force and is between zero and one. The second term is *β*, which is a first-order approximation of the life-saving (mortality-reducing) effect of healthcare providers on the probability of a patient dying from disease or disease-related complications. The third term is *y = f(c)/N*, which is the reduction in cases associated with a school closure.

The first term, *g(c)*, is rarely calculated. Lempel, Epstein, and Hammond provide a preliminary estimate for school closures in the US of a 6-19% reduction in the healthcare workforce.^1^ To provide detailed estimates, we use data from the US Current Population Survey (CPS) to quantify the impact of school closures on healthcare labor supply, *g(c)*. The CPS is an ongoing monthly survey of approximately 60,000 US households jointly administered by the US Census Bureau and Bureau of Labor Statistics. We access the data via the Integrated Public Use Microdata.^10^ We use the recent dataset, which includes information on just over 3 million individuals spread across 1.25 million households between January 2018-2020. The detailed data in the CPS allows us to characterize the family structure and likely within-household childcare options for healthcare workers. It also enables us to describe the exposure to child care obligations for specific occupations within healthcare and across states. We calculate the share of healthcare provider households that are single parent, where a parent is defined to include head of household, opposite or same-sex spouse, partner or roommate, and an opposite or same-sex unmarried partner. We use the personal CPS sample weights for all calculations to ensure that the estimates are nationally representative.

The second term, *β*, is calculated in development contexts and in emergency medicine, but to our knowledge has not been measured for accute infectious diseases (e.g., influenza or COVID-19) or included explicitly in epidemiological models. It is useful to know the critical value of *β* where school closure stops saving lives and starts increasing mortality, which is defined by the condition

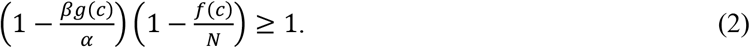

Imposing condition (2) as a strict equality and rearranging yields 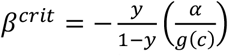, when *β* exceeds this value, then more lives are lost from school closures than saved. 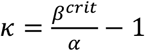 is the maximum percent increase in the mortality rate that does not reverse lives saved from school closures. *κ* is a more inutive quantity, and it also provides a number that is invariant to *α*.

The epidemiological literature has focused on the final term, *f(c)/N*. This value is usually determined by adjusting the conditional infectivity, either parametrically or through a behavioral model within an epidemiological model to account for a school closure.^11^ Models are required because there is little unconfounded experience with school closures during an epidemic, and few analyses of any behavior change are empirical.^12^ One of the most detailed studies is Cauchemez et al.,^2^ which estimates that prolonged school closures would reduce cumulative influenza cases by 13-17% in France, which implies *f(c)/N* is between 0.13 and 0.17. We focus on the midpoint of this estimate, 0.15, but consider cumlative case reductions from school closures between 0 and 50%.

## Results

Twenty-nine percent of healthcare worker households have an obligation to care for a child between 3 and 12 years of age. Focusing on households without a non-working adult or sibling 13 years or older who could potentially care for children ages 3-12 (https://www.redcross.org/take-a-class/babysitting/babysitting-child-care-training/babysitting-certification), we find that 15% or slightly more than 1 in 7, healthcare worker households have child care obligations. This may be optimistic because non-working adults may not be able to provide care, e.g., they may require care themselves. On the other hand, it is possible that family members outside the household, neighbors, or friends could care for children, though no data are available in the CPS on these possibilities. We find that 7% of healthcare households are single-parent households, which is greater than all other major industry classifications.

Within the healthcare sector, there are critical professions that are even more exposed to child care obligations (Table 1). These include households with a nurse, psychiatric, and home health care aids, nurse practitioner, medical assistant, nurse anesthetist, diagnostics technologist, physician’s assistant, or physician and surgeon. Assuming that non-working adults or siblings 13 or older can meet the child care obligations, 22% of nurse practitioners, 21% of physician’s assistants, 19% of diagnostic technicians, 18% of medical assistants, 16% of physicians and surgeons, and 13% of nursing, psychiatric, and home healthcare aids would still have unmet child care obligations. This is just over 20% of the healthcare workforce.

**Table 1.**
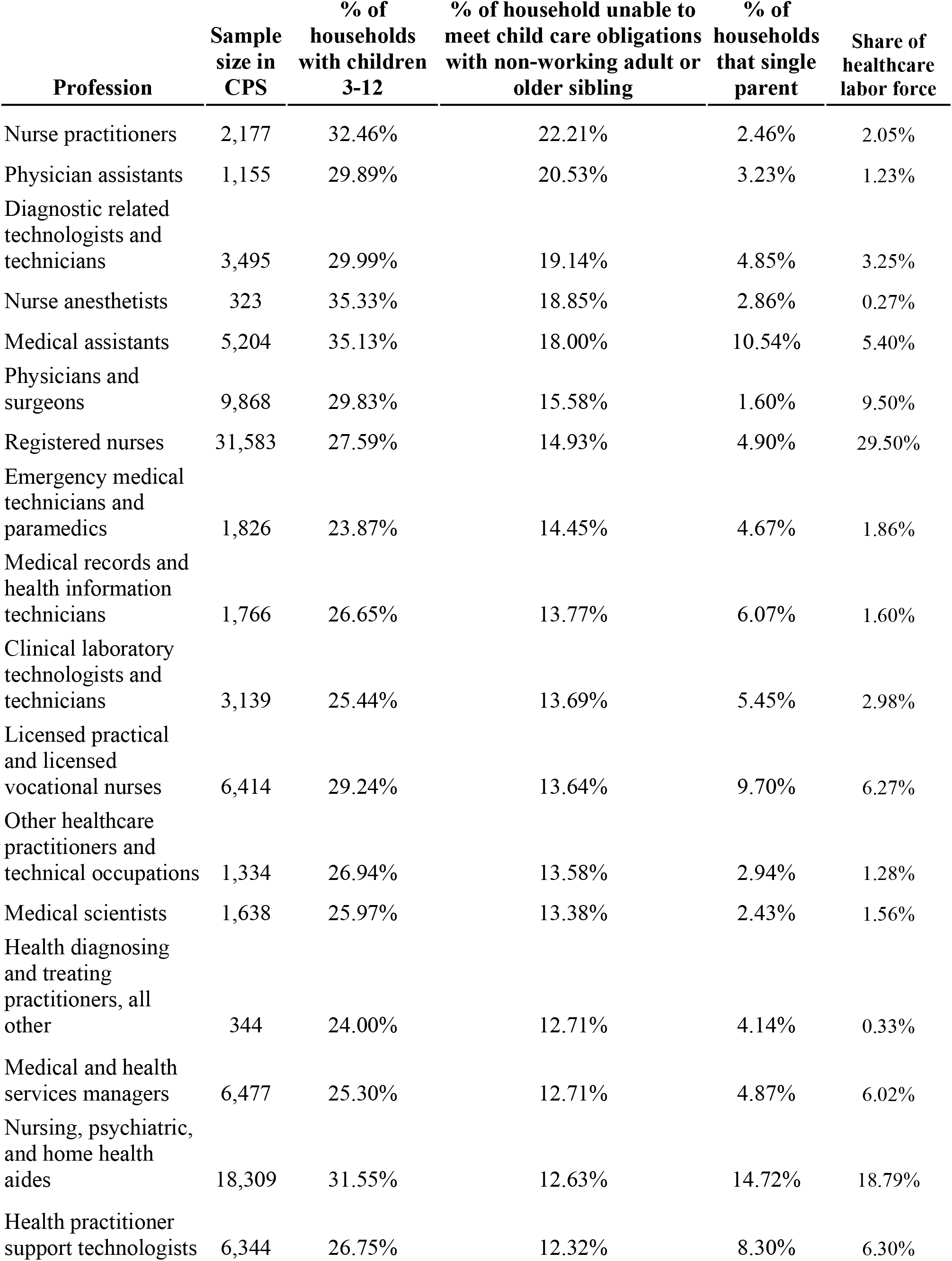
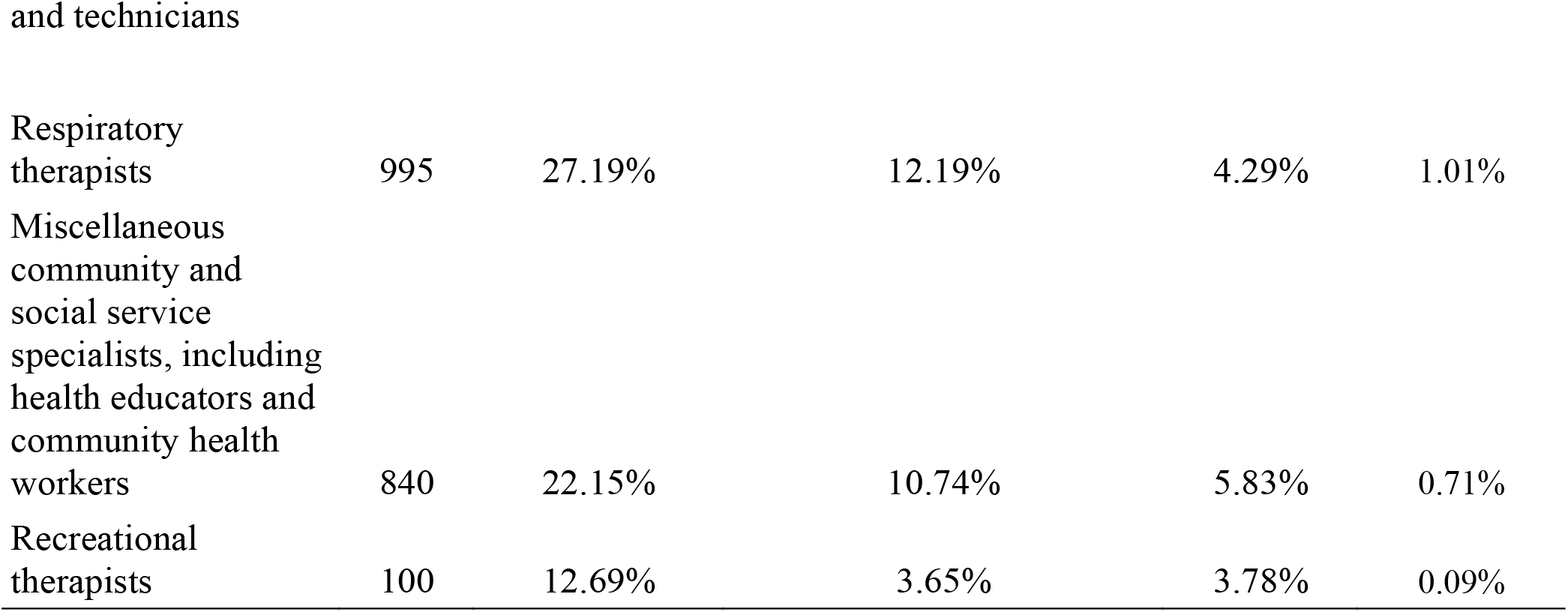
Child care obligations by healthcare profession

| **Profession** | **Sample size in CPS** | **% of households with children 3-12** | **% of household unable to meet child care obligations with non-working adult or older sibling** | **% of households that single parent** | **Share of healthcare labor force** |
| --- | --- | --- | --- | --- | --- |
| Nurse practitioners | 2,177 | 32.46% | 22.21% | 2.46% | 2.05% |
| Physician assistants | 1,155 | 29.89% | 20.53% | 3.23% | 1.23% |
| Diagnostic related technologists and technicians | 3,495 | 29.99% | 19.14% | 4.85% | 3.25% |
| Nurse anesthetists | 323 | 35.33% | 18.85% | 2.86% | 0.27% |
| Medical assistants | 5,204 | 35.13% | 18.00% | 10.54% | 5.40% |
| Physicians and surgeons | 9,868 | 29.83% | 15.58% | 1.60% | 9.50% |
| Registered nurses | 31,583 | 27.59% | 14.93% | 4.90% | 29.50% |
| Emergency medical technicians and paramedics | 1,826 | 23.87% | 14.45% | 4.67% | 1.86% |
| Medical records and health information technicians | 1,766 | 26.65% | 13.77% | 6.07% | 1.60% |
| Clinical laboratory technologists and technicians | 3,139 | 25.44% | 13.69% | 5.45% | 2.98% |
| Licensed practical and licensed vocational nurses | 6,414 | 29.24% | 13.64% | 9.70% | 6.27% |
| Other healthcare practitioners and technical occupations | 1,334 | 26.94% | 13.58% | 2.94% | 1.28% |
| Medical scientists | 1,638 | 25.97% | 13.38% | 2.43% | 1.56% |
| Health diagnosing and treating practitioners, all other | 344 | 24.00% | 12.71% | 4.14% | 0.33% |
| Medical and health services managers | 6,477 | 25.30% | 12.71% | 4.87% | 6.02% |
| Nursing, psychiatric, and home health aides | 18,309 | 31.55% | 12.63% | 14.72% | 18.79% |
| Health practitioner support technologists | 6,344 | 26.75% | 12.32% | 8.30% | 6.30% |
| Respiratory therapists | 995 | 27.19% | 12.19% | 4.29% | 1.01% |
| Miscellaneous community and social service specialists, including health educators and community health workers | 840 | 22.15% | 10.74% | 5.83% | 0.71% |
| Recreational therapists | 100 | 12.69% | 3.65% | 3.78% | 0.09% |

School closures may be especially challenging for single parents. 14.7% of nursing, psychiatric, and home health aides are single parents. Medical assistants (10.5%) and licensed practical and licensed vocational nurses (9.7%) are also more likely to be single parents than the healthcare average. Together this is 30% of the healthcare workforce, and is the segment most likely to be providing infection control for the elderly in nursing homes and other facilities.

Registered nursing is the most common profession in the healthcare field and accounts for 29.5% of the healthcare labor force, followed by nursing, psychiatric, and home health care aides (18.8%). 28% of registered nurses have child care obligations, but perhaps 13% of registered nurses can meet these obligations with a non-working adult or older sibling, leaving 15% with unmet child care obligations. Five percent of registered nurses are single parents, but this varies from state to state (Figure 1).

**Figure 1.**
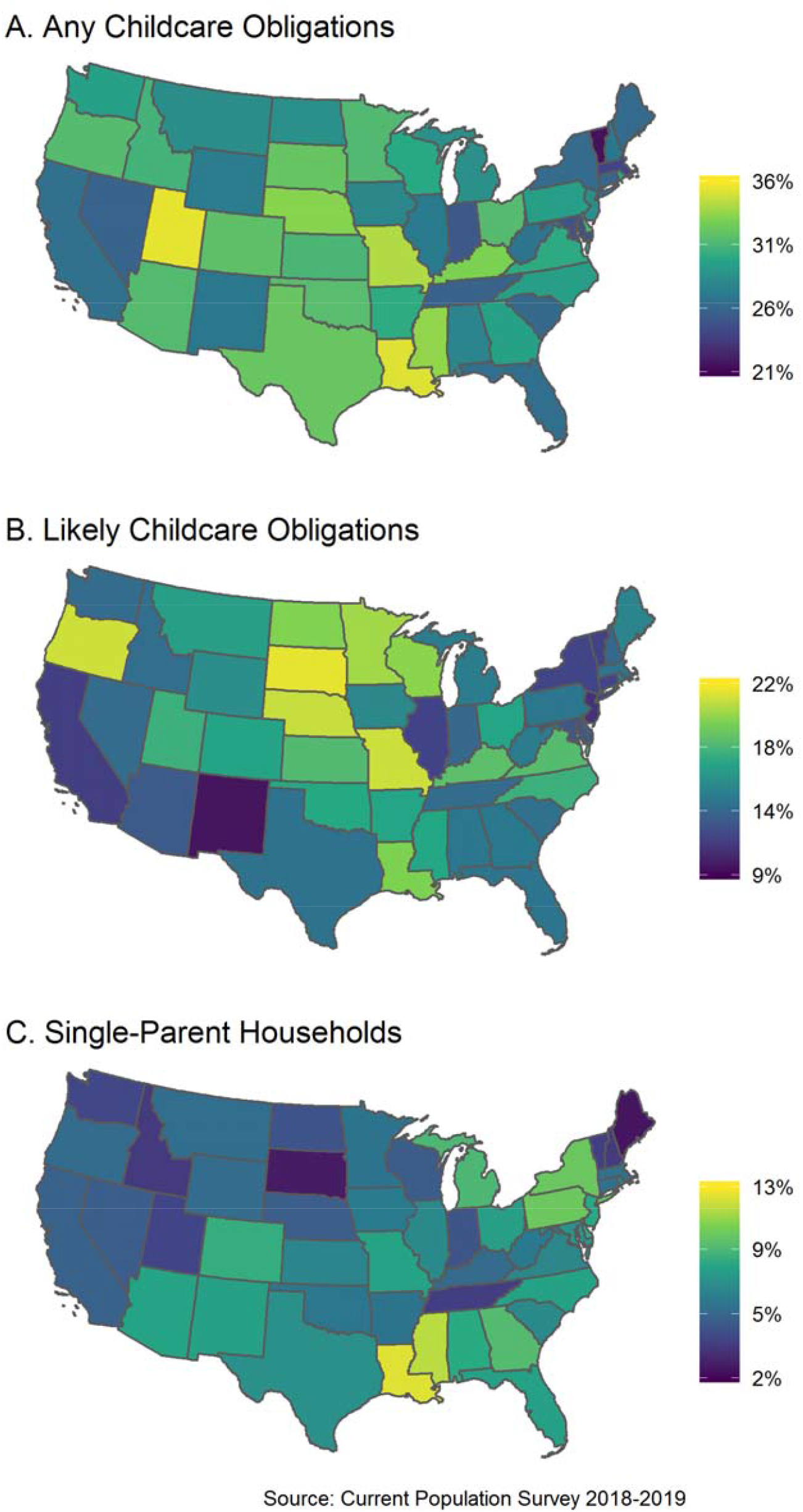
The map depicts the fraction of the healthcare workforce with possible childcare obligations under various adaptation assumptions: A) healthcare workers in households with at least one child age 3-12, B) heathcare workers in households with at least one child age 3-12 and without a non-working adult or child over 12 that might provide childcare, and C) healthcare workers in single-parent households.

The exposure of the healthcare labor force to school closures is not homogeneous throughout the US (Appendix Table 1). How a policy trade-off plays out in one place may be different than another even if two locations experience a similar set of COVID-19 cases. That is, different places may have similar projected benefits from school closures, but different costs. The greatest share of the healthcare labor force with child care obligations is in Utah (35%), Louisiana (35%), and Missouri (34%), whereas the healthcare labor force in Washington DC (16%), Vermont (21%), and Massachusetts (24%) have the least child care obligations. However, household structures also vary. Therefore, if childcare obligations can be met by a non-working adult or older sibling, then South Dakota (21%), Oregon (20%), and Missouri (20%) are the most exposed states to school closure induced healthcare worker shortages. Washington DC (9%), New Mexico (10%), and New Jersey (11%) may be most able to cover their child care obligations. Louisiana (12%), Mississippi (11%), and Pennsylvania (10%) have the greatest fraction of single-parent healthcare worker households. These differences are likely an interaction of variations in state-level healthcare regulation, cultural, and demographic differences. This requires state and local health officials to consider the exposure of their own state (data available at https://github.com/jbayham/us_childcare_obligations) or region.

Using the data on child care obligations provides a first estimate of potential reductions in the healthcare labor force during a school closure. This estimate can be combined with projections of the case reductions from school closures to identify the parameter space where estimates of 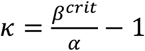 are important to inform whether school closures would reduce net mortality (Figure 2). If the *κ* is large, then the increase percent increase in the mortality rate is below *κ*, and closing schools saves lives. This is the case when school closures lead to many avoided cases, and few healthcare workforce effects. Conversely, if *κ* is near zero, then then it is likely that the percent increase in mortality exceeds *κ* and school closure increases the cumulative mortality. This is the case if the school closure reduces the labor force substantially but provides a relatively small reduction in cumulative cases. There is a band where *0 ≪ κ ≪ ∞*, where whether or not closing schools saves lives depends on the value of *β*, which is unknown.

**Figure 2.**
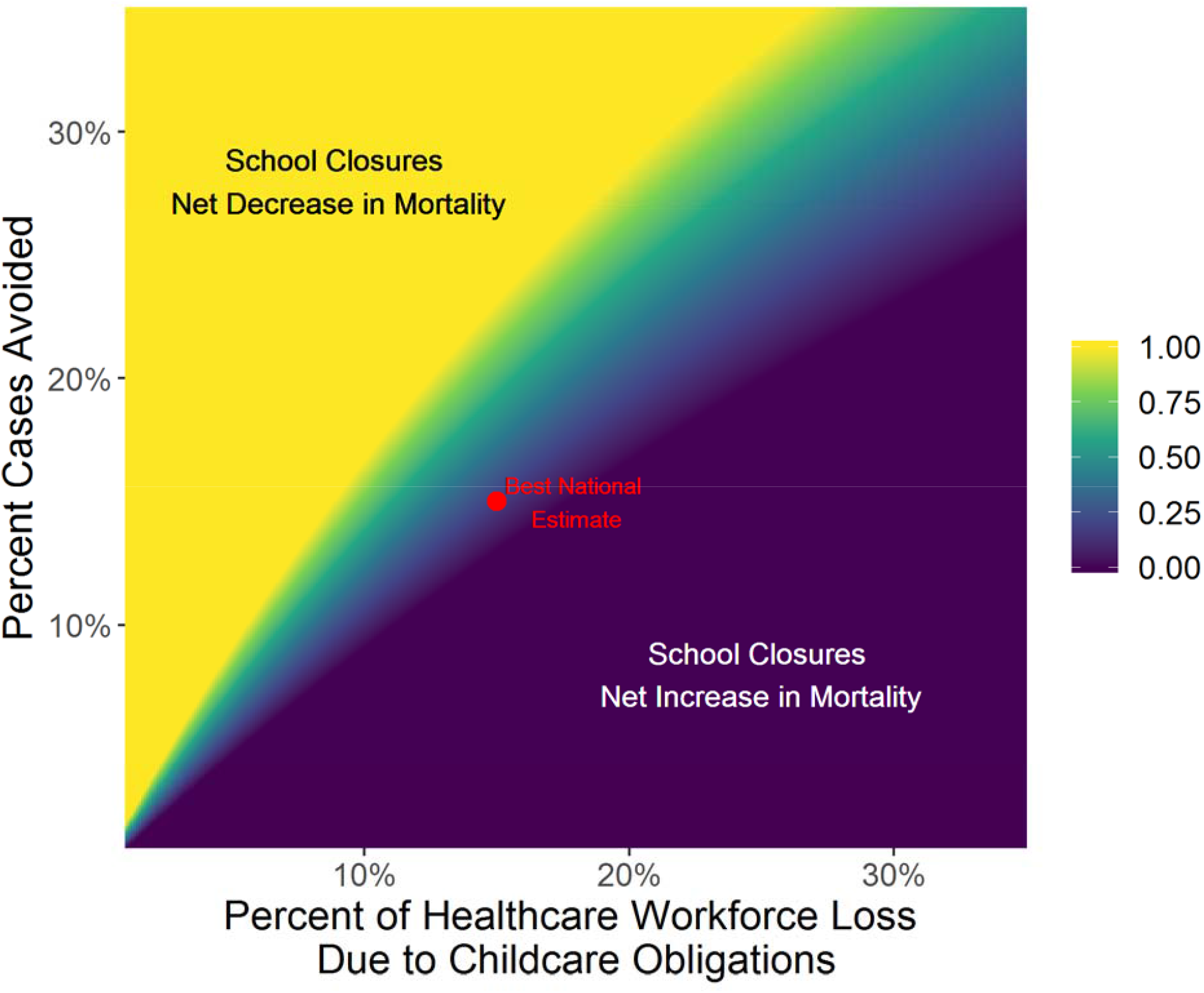
Critical level of the percent increase in mortality resulting from healthcare work force absenteeism associated with school closure induced child care obligations,*κ*, that would offset the mortality reduction achieved by school closures through case reductions (color scale). The actual percent increase in mortality must be below *κ* to justify closing schools. The red point, *κ* = 0.176, indicates the best national estimates of cases avoided due to school closures (15% [13%,17%]) and the mean estimates of unmet healthcare workforce childcare obligations, 15%. This estimate accounts for the potential for other non-working adults or older siblings in the household to provide childcare.

For the United States, and for most states within the US, the *κ* is not sufficently higher or lower to know which way a school closure will turnout without more information on *β*. Consider an example that uses Cauchemez et al.’s (2008) estimate of a reduciton of cases of 15% from a school closure and assume baseline mortality to COVID-19 of 2%. Assume an epidemiological forcast without a school closure predicts 16,000 cases (about a 1/5^th^ of the cases currently reported in China), which implies 320 deaths (Italy has already reported 463 deaths with 9,172 cases). The school closure reduces cases to 13,600. The *κ* for this scenario is 0.176, with an associate *β*^*crit*^ = 0.024. Therefore, the mortality rate after the closure must rise to at least 2.35% as a result of the 15% loss in the healthcare workforce in order to undo the benefits of school closures. This implies the percent increase in patient survival by avoiding a 15% reduction in the health workforce, an elasticity measuring healthcare worker “prodcutivity,” is 0.024. This means that doubling the healthcare workforce must not save more than 2.4% more patients or school closures could lead to more deaths. However, there is substantial variation across the country. For example, in South Dakota this elasticity is 1.7%, wheres in Washington DC it is 4.1%.

An additional concern could be timing within an epidemic. School closure could spread out cases so as not to overwhelm the healthcare system. Cauchemez et al. estimates a reduction in peak prevalence of about 42%. This cannot raise the mortality per case in that period to less than 3.4% or an elasiticity of 9.9% for school closure to save lives.

## Discussion

What we know about social distancing policies is based largely on models of influenza,^4,13^ where children are a vulnerable group. However, preliminary data on COVID-19 suggests that children are a small fraction of cases and may be less vulnerable than older adults.^14^ If these early results hold-up, then the already uncertain transmission reduction benefits from school closures will be reduced relative to influenza. Conversely, school closures may be implemented earlier in COVID-19 outbreaks, which may lead to greater levels of prevented cases. Furthermore, school closures may lead to other adults staying home, which could reduce cases. These are all important questions when considering school closures.

School closures do expose the healthcare labor force to increased child care obligations, likely reducing support for infected individuals, which is critical. We also do not know how much a reduction in the healthcare labor force, and in what occupations within healthcare, decreases the probability of COVID-19 patient survival. We do know that the segment of the healthcare work force most responsible for infection control in nursing homes is likely to be among the most highly impacted by school closure induced child care obligations. Given our reasonable estimates of case reductions from school closures, a measure of the increased mortality risk to COVID-19 patients from healthcare absenteeism to care for children is a critical, and to date unknown parameter. This analysis does not include non-COVID-19 mortality that could occur from other conditions if the US healthcare work force is reduced, but risk to these patients should also be considered when deciding about school closings.

Minimizing the impact of the COVID-19 and saving lives requires clear thinking and weighing tradeoffs. There will likely be cases when closing schools is sensible. However, policymakers and the healthcare experts advising them need to understand that closing schools may have severe knock-on effects on the healthcare system and that there is substantial uncertainty about the effectiveness of school closures for preventing infection beyond school children and on the impact of reduction in the healthcare workforce on patient survival. In the United States, the healthcare system appears disproportionately exposed to school closure induced labor shortages, and the segment of that system that provides infection control in nursing homes even more so. Such potential healthcare workforce labor shortages should be a first-order consideration when considering the potential benefits and costs of school closures.

## Data Availability

The US Current Population Survey is publically available, see Flood et al. Code to access and organize the data are hosted on Bayham’s Github page, https://github.com/jbayham/us_childcare_obligations.

## Author Contributions

Both authors worked closely to design and write the manuscript. JB lead the data management and analysis.

---

**Appendix 1.**
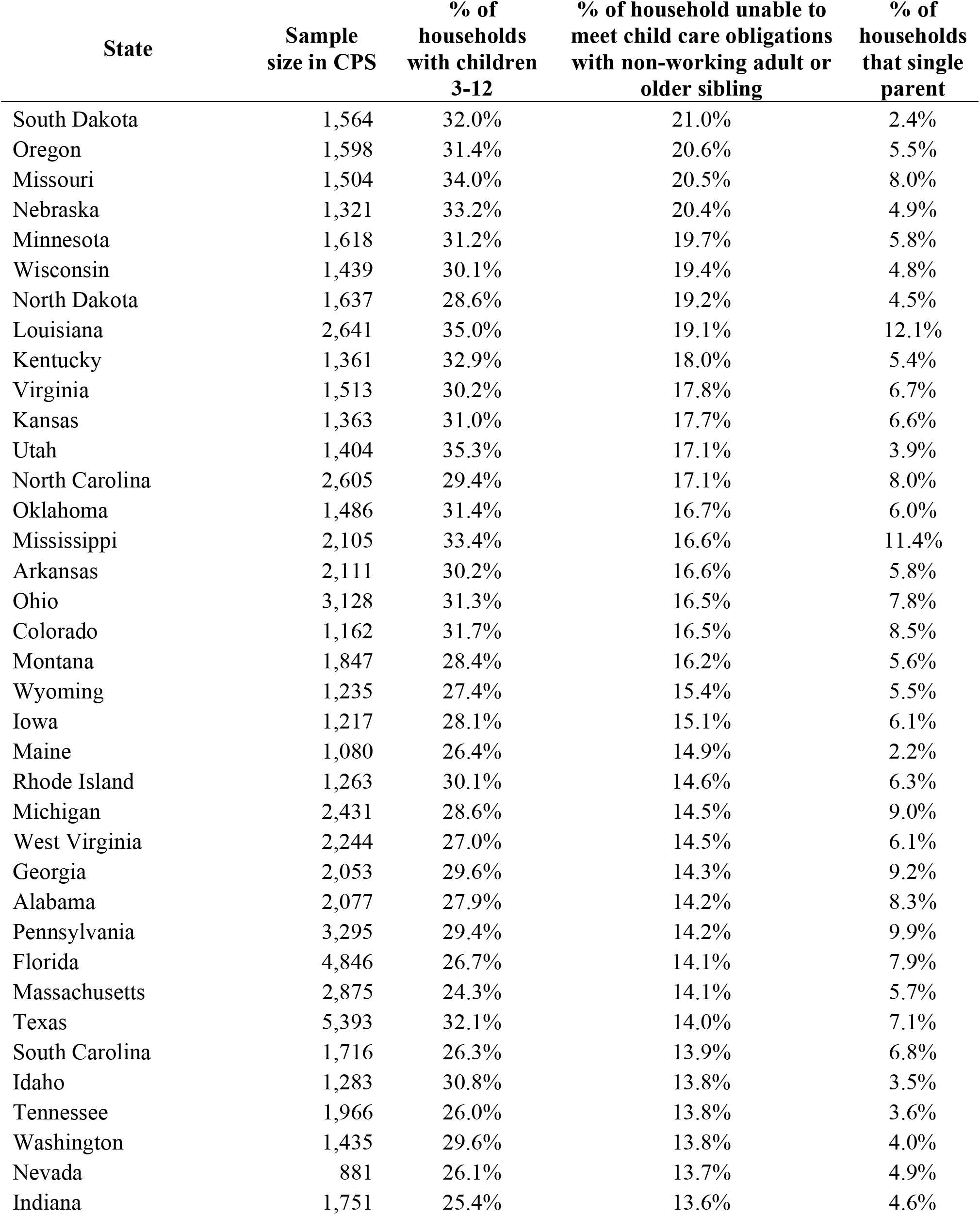
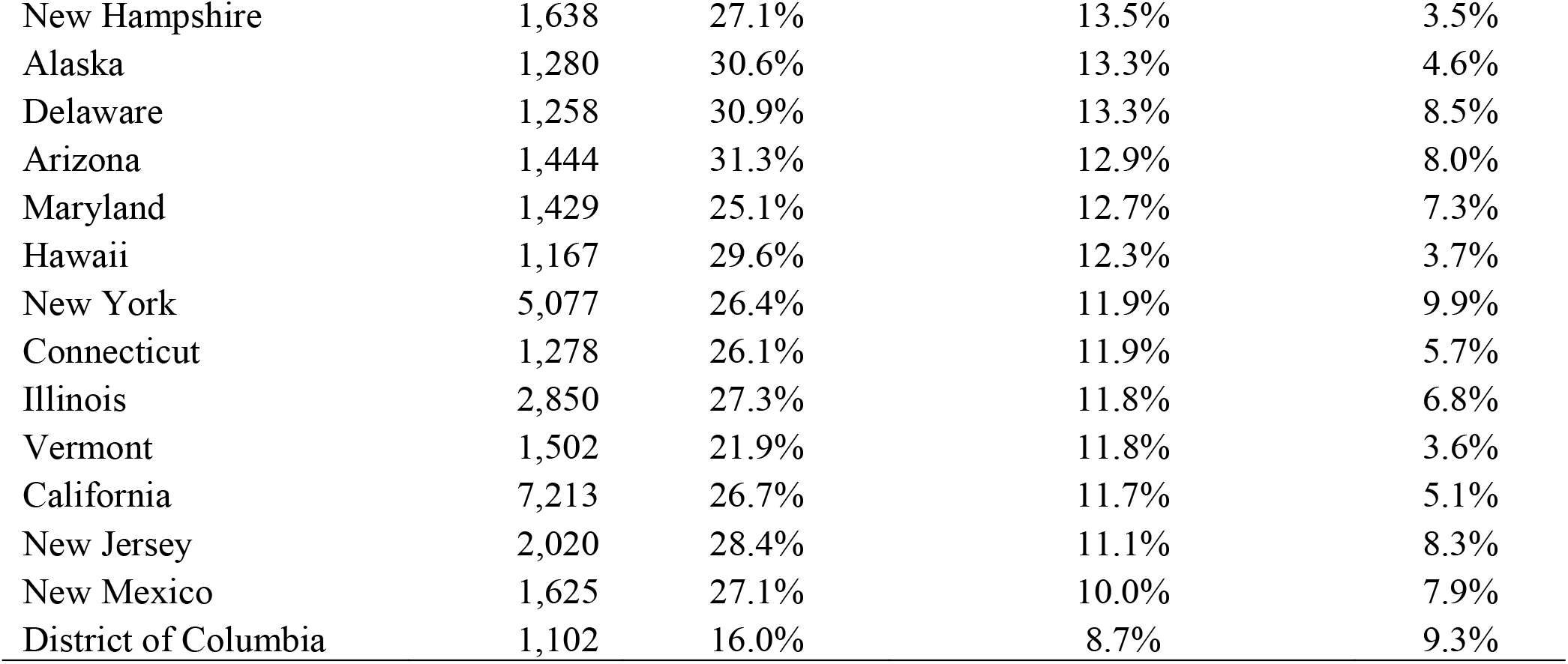
Healthcare worker child care obligations by state, including Washington DC, ranked by unmet childcare obligations after allowing for non-working adults or children over 12 to care for younger children in the household.

